# First Clinical Study Using HCV Protease Inhibitor Danoprevir to Treat Naïve and Experienced COVID-19 Patients

**DOI:** 10.1101/2020.03.22.20034041

**Authors:** Hongyi Chen, Zhicheng Zhang, Li Wang, Zhihua Huang, Fanghua Gong, Xiaodong Li, Yahong Chen, Jinzi J. Wu

## Abstract

As coronavirus disease 2019 (COVID-19) outbreak, caused by the severe acute respiratory syndrome coronavirus-2 (SARS-CoV-2), started in China in January, 2020, repurposing approved drugs is emerging as important therapeutic options. We reported here the first clinical study using hepatitis C virus (HCV) protease inhibitor, danoprevir, to treat COVID-19 patients. Danoprevir (Ganovo®) is a potent HCV protease (NS3/4A) inhibitor (IC50 = 0.29 nM), which was approved and marketed in China since 2018 to treat chronic hepatitis C patients. Ritonavir is a CYP3A4 inhibitor to enhance plasma concentration of danoprevir while it also acts as a human immunodeficiency virus (HIV) protease inhibitor at high doses. The chymotrypsin-like protease of SARS-CoV-2 shares structure similarity with HCV and HIV proteases. In the current clinical study (NCT04291729) conducted at the Nineth Hospital of Nanchang, we evaluated therapeutic effects of danoprevir, boosted by ritonavir, on treatment naïve and experienced COVID-19 patients. The data from this small-sample clinical study showed that danoprevir boosted by ritonavir is safe and well tolerated in all patients. After 4 to 12-day treatment of danoprevir boosted by ritonavir, all eleven patients enrolled, two naïve and nine experienced, were discharged from the hospital as they met all four conditions as follows: (1) normal body temperature for at least 3 days; (2) significantly improved respiratory symptoms; (3) lung imaging shows obvious absorption and recovery of acute exudative lesion; and (4) two consecutive RT-PCR negative tests of SARS-CoV-2 nucleotide acid (respiratory track sampling with interval at least one day). Our findings suggest that repurposing danoprevir for COVID-19 is a promising therapeutic option.

## Introduction

The severe acute respiratory syndrome coronavirus-2 (SARS-CoV-2) has spread rapidly since its identification in patients with severe pneumonia in Wuhan, China. The Coronavirus disease-19 (COVID-19), the disease caused by SARS-CoV-2 infection, has been reported more than 118,000 cases in 114 countries, and 4,291 people have lost their lives by March 11, 2020 [1]. Given its global outbreak and strong human-to-human transmission and high mortality in elderly or in patients with complications, the World Health Organization (WHO) has made the assessment on March 11, 2020 that COVID-19 can be characterized as a pandemic [2].

Investigation into approved drugs for new therapeutic indications, which is also known as drug repurposing, has been proved to be a practical strategy in the situation of an outbreak such as COVID-19 [3-4]. Danoprevir is a potent hepatitis C virus (HCV) protease inhibitor with an IC_50_ of 0.29 nM against HCV protease [5] and was approved and marketed in 2018 in China as an oral direct-acting antiviral agent to treat hepatitis C. The results of a phase 3 clinical trial with 140 patients showed that the triple regimen of ritonavir-boosted danoprevir plus pegylated-interferon a-2a and ribavirin produced a sustained virologic response (SVR12) rate of 97.1% after 12-week treatment in non-cirrhotic hepatitis C virus (HCV) genotype 1-infected Chinese patients [6]. In another phase 2/3 clinical trial, the SVR12 rate is 99% in treatment-naïve, non-cirrhotic HCV GT1-infected patients after 12-week all oral treatment of ritonavir-boosted danoprevir combined with ravidasvir (an HCV NS5A inhibitor) and ribavirin [7]. These results indicated danoprevir boosted by ritonavir is highly efficacious, safe, and well tolerated in HCV patients.

During SARS-COV-2’s life cycle, chymotrypsin-like protease (3CL pro) can cleave viral polyprotein to form the RNA replicase-transcriptase complex, which is essential for both viral transcription and replication [8-9]. The proteases of HCV and human immunodeficiency virus (HIV) showed similar function as SARS-COV-2, so protease inhibitors are hypothesised to have the therapeutic potential against COVID-19. *In vitro* and clinical studies in patients with SARS and middle east respiratory syndrome (MERS) to some extent proved this hypothesis [10-12]. The homology modeling data indicated that HCV protease inhibitors have the highest binding affinity to SARS-CoV-2 protease among approved drugs [13]. Yet there is still a paucity of clinical data on HCV protease inhibitors in treating COVID-19 patients.

In this study, we for the first time demonstrated the therapeutic efficacy of danoprevir boosted by ritonavir in both treatment naïve and experienced COVID-19 patients.

## Methods

### Diagnosis

According to the “Diagnosis and Treatment Program for Novel Coronavirus Infection (Trial Version 6)” issued by the National Health Commission of the People’s Republic of China (China National Standards) [14], patients with RT-PCR positive of SARS-CoV-2 nucleotide acid are diagnosed as SARS-CoV-2 infection. COVID-19 patients, demonstrating pneumonia manifestation in imaging, respiratory tract symptoms and RT-PCR positive of SARS-CoV-2 nucleotide acid, are categorized as moderate COVID-19 patients per China National Standards [14].

### Study design, patients, treatments and discharge

In this open clinical trial for the treatment of COVID-19 (ClinicalTrials.gov Identifier: NCT04291729), patients were treated with danoprevir boosted by ritonavir in the presence or absence of interferon nebulization (the background therapy). 10 patients were planned to be enrolled and actually 11 patients were enrolled. All patients were moderate COVID-19 patients per China National Standards [14]. These patients were recruited and screened, and eligible patients were enrolled and treated with danoprevir boosted by ritonavir in the presence or absence of interferon nebulization (as the background therapy) for 4 to12 days until being discharged from the hospital. Treatment naïve patients never received any antiviral therapies such as lopinavir/ritonavir and interferon nebulization prior to the treatment of danoprevir boosted by ritonavir. Treatment experienced patients had received at least one dose of any antiviral therapies such as lopinavir/ritonavir and interferon nebulization prior to the treatment of danoprevir boosted by ritonavir. Treatment regimen and dosing frequency are: (1) danoprevir, 100 mg per tablet, 1 tablet at a time, twice per day; (2) ritonavir, 100 mg per tablet, 1 tablet at a time, twice per day; (3) if administered, aerosol inhalation of α-interferon (interferon nebulization), 5 million units at a time, twice per day.

The conditions of the patients were monitored daily by attending physicians. Routine laboratory test of blood indexes, biochemical parameters, and chest computer tomography (CT) were carried out in need.

According to China National Standards [14], the following four conditions must be all met prior to the discharge: (1) normal body temperature for more than 3 days; (2) significantly recovered respiratory symptoms; (3) lung imaging shows obvious absorption and recovery of acute exudative lesion; and (4) two consecutive RT-PCR negative tests of SARS-CoV-2 nucleotide acid (respiratory track sampling with interval at least one day). Furthermore, patients were reviewed by expert committee comprised of elite clinicians before the discharge.

All patients signed informed consent form. The Ethics Committee of the hospital approved the study.

### Inclusion Criteria

Chinese adult (18-75 years old in both genders) pneumonia patients with SARS-COV-2 infection confirmed to be positive by RT-PCR; demonstrated respiratory symptoms and imaging-confirmed pneumonia as defined in China National Standards; naïve patients were newly diagnosed patients who never received any antiviral treatments and danoprevir/ritonavir treatment were initiated within 7 days or less from the diagnosis; experienced patients might receive antiviral therapies with dosage and treatment duration recommended by China National Standards [14]; women and their partners with no planned pregnancy for 6 months after the study and willing to take effective contraceptive measures within 30 days from the first administration of the study drugs; patients agreed that they would not participate in other clinical trials within 30 days from the last administration of the study drugs; patients were willing to sign informed consent form.

### Exclusion Criteria

Severe COVID-19 patients met one of the following conditions: (1) Respiratory rate (RR) ≥ 30 times / min; (2) SaO2 / SpO2 ≤ 93% in resting state; (3) arterial partial pressure of oxygen (PaO2) / concentration of oxygen (FiO2) ≤ 300 mmHg (1mmhg = 0.133 kPa; critical COVID-19 patients met one of the following conditions: (1) respiratory failure and need mechanical ventilation; (2) shock; (3) other organ failure combined with ICU treatment; severe liver disease (such as child Pugh score ≥ C, AST > 5 times upper limit); patients with contraindications specified in the label of ritonavir tablets; the pregnancy test of female subjects during the screening period was positive; the researchers judged that the patient was not suitable to participate in this clinical study (for example, patients with multiple basic diseases, etc.).

### RT-PCR protocol

Real-time PCR (RT-PCR) testing of SARS-COV-2 ORF1ab gene was performed in the clinical laboratory of the Ninth Hospital of Nanchang following the test kit instruction of DAAN Gene (manufacturer), Guangzhou, China. The lowest detection limit of this kit is 1×10^3^ copies/ml. Briefly, Nasal and phlegm swab specimens were collected and then put in a sterile tube containing 2 to 3 ml of viral transport medium. Total cellular RNA was isolated by TRIzol reagent (Invitrogen) from 200 µl transport medium. RT-PCR was conducted in LightCycler 480 (Roche) by mixing 17 µl PCR solution A, 3 µl PCR solution B and 5 µl sample RNA in one tube. A positive control was used to determine the positivity of the samples.

### CT scans

Chest CT scans were conducted using Highspeed dual-slice spiral CT (GE, USA). All CT images were axial thin-section non-contrast and were undergone 5 mm reconstruction.

### Efficacy endpoints

Efficacy endpoints include (1) time to discharge; (2) rate of two consecutive RT-PCR negative tests of SARS-CoV-2 nucleotide acid (respiratory track sampling and sampling interval at least one day apart); (3) lung imaging showing obvious absorption and recovery of acute exudative lesion ; (4) significantly improved respiratory symptoms; (5) normal body temperature for at least 3 days.

## Results

### Baseline demographic and clinical characteristics

A total of 11 moderate COVID-19 patients were enrolled, including two naive patients and nine experienced patients. The baseline demographic characteristics for all enrolled patients are illustrated in Table 1. The age range of the patients were 18 to 66 years old, with 4 males in total 11 patients. Two experienced patients, patient <u>6</u> and patient <u>8</u>, had a history of hypertension. The symptoms reported at illness onset were mainly fever, cough and shortness of breath. Two naive patients were hospitalized for 9 and 7 days, respectively, and the experienced patients were hospitalized for a median duration of 20 days.

**Table 1.** Baseline demographic characteristics, underlying medical condition, and clinical characteristics of 11 patients receiving danoprevir boosted by ritonavir treatment

|  | Naive<br>(N=2) | Experienced<br>(N=9) |
| --- | --- | --- |
| Median age – years | 44, 18 | 44 (18–66)<sup>#</sup> |
| Median hospitalization – days | 9, 7 | 20 (7–22)<sup>#</sup> |
| Male | 1 | 3 |
| Underlying medical conditions | no, no | 2<sup>\$</sup> |
| Symptoms reported at illness onset |  |  |
| Fever | no, no | 4 |
| Cough | no, yes | 3 |
| Shortness of breath | no, yes | 6 |

<sup>#</sup> The values were presented as median with maximum and minimum values in bracket.
<sup>\$</sup> Two experienced patients, patient ~~5~~6 and patient ~~7~~8, had a history of hypertension.

The key parameters of 11 patients were presented in Table 2 and illustrated graphically in Figure 1. In general, nine patients experienced lopinavir/ritonavir along with interferon nebulization treatment were enrolled and switched to danoprevir/ritonavir treatment, including four patients with interferon nebulization and five patients without interferon nebulization after switching. Two naive patients received danoprevir/ritonavir plus interferon nebulization treatment. After initiation of danoprevir/ritonavir treatment, the first negative RT-PCR test occurred at a median of 2 days, ranging from 1 to 8 days, and the obvious absorption in CT scans occurred at a median 3 days, ranging from 2 to 4 days.

**Table 2.** Key parameters of 11 patients receiving danopreivr boosted by ritonavir treatment

|  | Days of hospitalization | Antiviral experience before danoprevir/ritonavir treatment | Days of lopinavir/ritonavir plus IFN nebulization treatment prior to the study | Days of danoprevir/ritonavir plus IFN nebulization treatment during the study | Days of danoprevir/ritonavir treatment without IFN nebulization during the study | Days of first negative RT-PCR test after initiation of danoprevir/ritonavir treatment* | Days of obvious absorption in CT scans after initiation of danoprevir/ritonavir treatment |
| --- | --- | --- | --- | --- | --- | --- | --- |
| Patient 1 | 9 | Naive | NA | 9 | NA | 2 | 4 |
| Patient 2 | 7 | Naive | NA | 7 | NA | 4 | 2 |
| Patient 3 | 7 | Experienced | 3 | 4 | NA | 1 | 2 |
| Patient 4 | 10 | Experienced | 2 | 8 | NA | 1 | 4 |
| Patient 5 | 17 | Experienced | 5 | 12 | NA | 2 | 3 |
| Patient 6 | 9 | Experienced | 3 | 6 | NA | 3 | 3 |
| Patient 7 | 22 | Experienced | 13 | NA | 9 | 2 | 3 |
| Patient 8 | 20 | Experienced | 11 | NA | 5 | 2 | 3 |
| Patient 9 | 21 | Experienced | 13 | NA | 8 | 2 | 4 |
| Patient 10 | 21 | Experienced | 13 | NA | 8 | 4 | # |
| Patient 11 | 22 | Experienced | 10 | NA | 9 | 8 | 3 |

NA, not applicable.
\* First negative tests were confirmed by second RT-PCR tests with at least 1-day interval.

**Figure 1.**
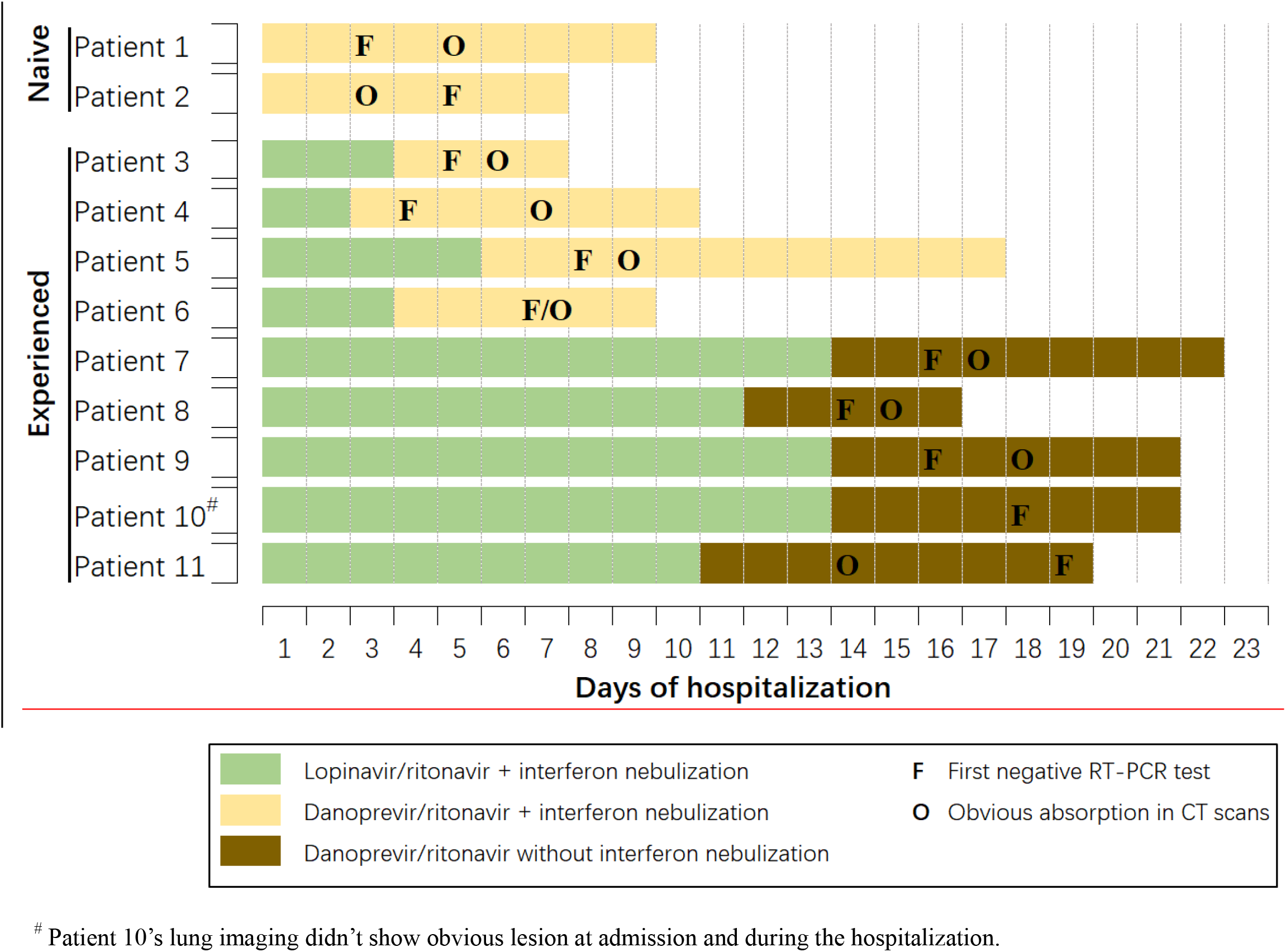
Timeline of RT-PCR tests, CT scans, and antiviral treatments of the enrolled patients.

### Naïve patients receiving danoprevir/ritonavir plus interferon nebulization

Two treatment naïve patients received danoprevir/ritonavir plus interferon nebulization upon hospitalization. The medical history, treatment and various examinations for one of two patients, patient 1, were described in details below.

- On February 19, 2020, a woman in her 40’s, who had a close contact with a COVID-19 patient, was tested positive SARS-COV-2 in nasal swab with RT-PCR and thus admitted to the hospital. The vital sign of the patient showed body temperature (BT) of 36.4°C and oxygen saturation (SPO_2_) of 99% under ambient air. The breathing sound was normal initially. She had no other underlying disease before this onset.
- In the morning of February 20, she was given the first dose of danoprevir boosted by ritonavir, along with interferon nebulization.
- On February 20, both lower lungs showed multiple diffuse patch-shape areas and ground-glass opacity (GGO), especially obvious in the left lower lung, based on chest CT scans (Figure 2A). The laboratory results revealed lymphopenia (0.62 ×10^9^ /L) and elevated platelet (322 ×10^9^ /L) (Table 3).
- On February 22, 3 days after receiving danoprevir boosted by ritonavir treatment, the patient didn’t show abnormal symptoms or vital signs. The lower lobe of bilateral lung showed decreased GGO and patch-shaped areas compared to the previous image on February 20, indicating partial absorption (Figure 2B). The RT-PCR results of nasal swab became negative. These results indicated the patient had a quick response to the treatment of danoprevir boosted by ritonavir, along with interferon nebulization.
- On February 24, 5 days after receiving danoprevir boosted by ritonavir treatment, the GGO and patch-shaped areas in both lungs significantly decreased in density and scale (Figure 2C) compared to the previous image on February 22. And the RT-PCR results of nasal swab were repeatedly negative on February24 and February 26.
- On February 28, Patient #1 was given the last dose of danoprevir boosted by ritonavir, along with interferon nebulization. Since she met all four discharge standards described in Methods section, the patient was discharged from the hospital on the same day.

**Table 3.**
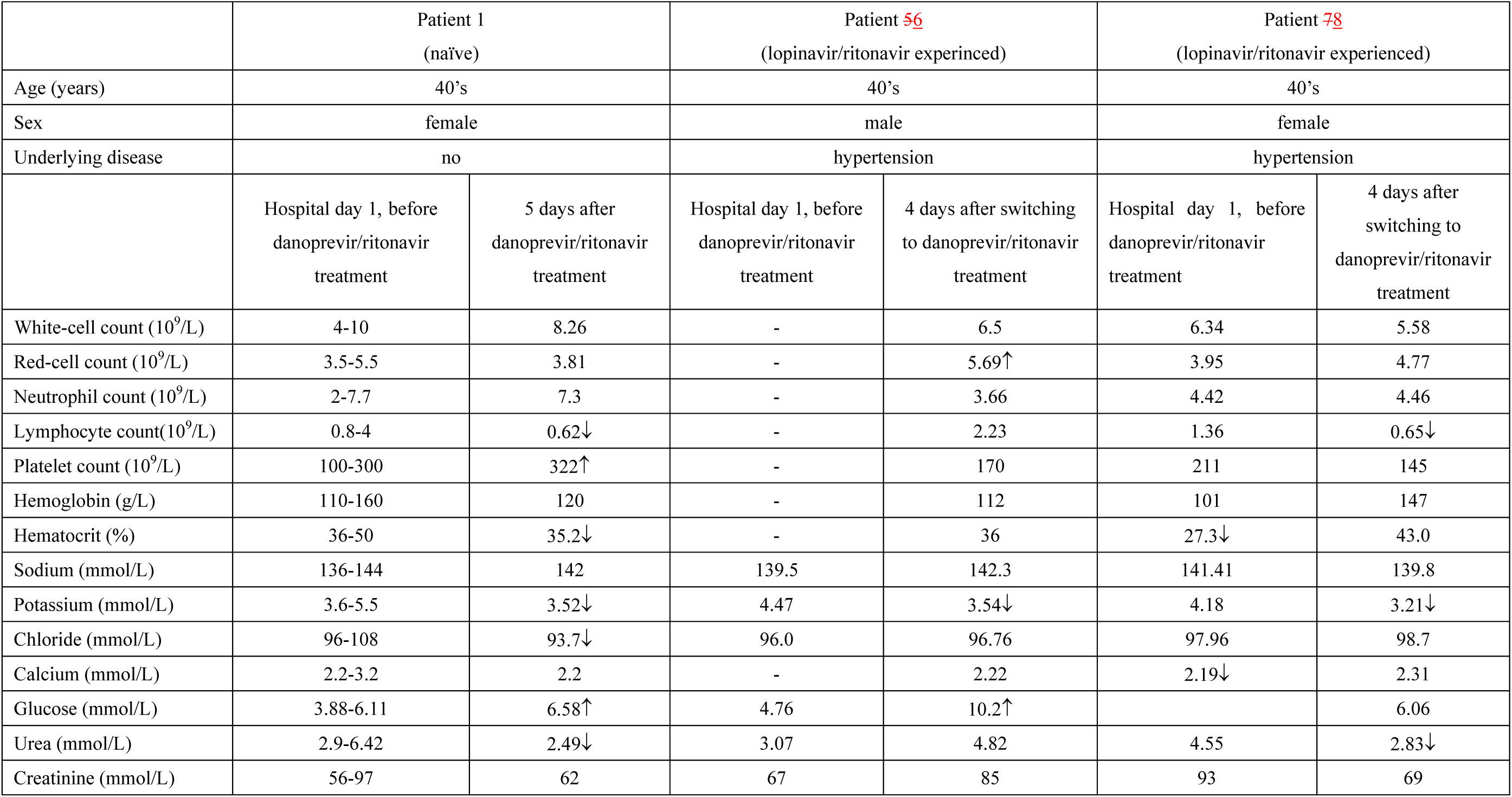
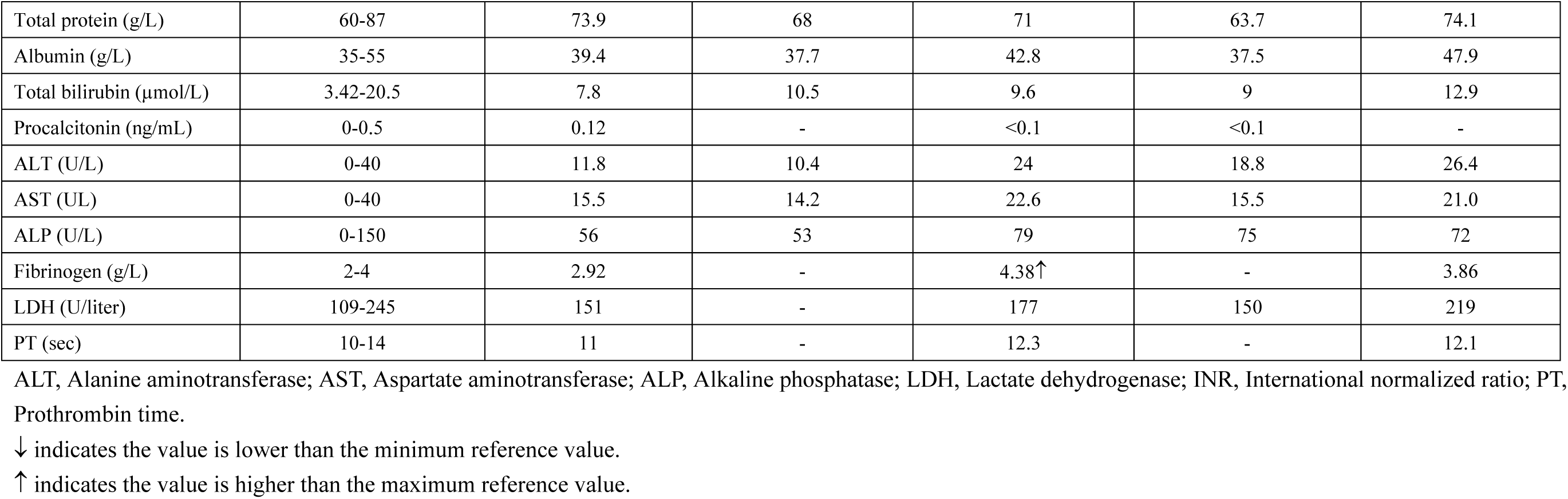
Laboratory tests and underlying disease of three representative patients before and after receiving danopreivr boosted by ritonavir treatment

**Figure 2.**
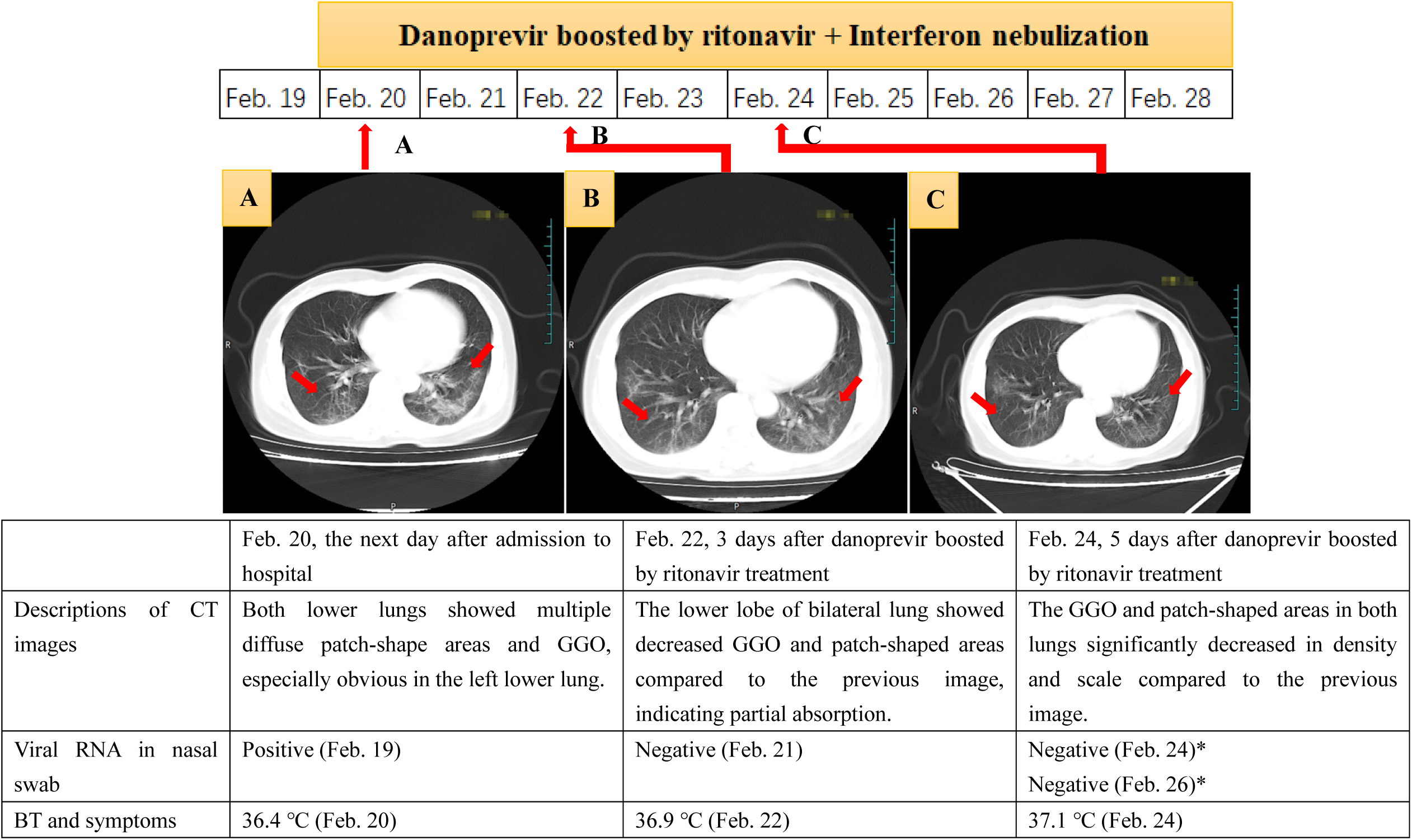
Naïve patient receiving danoprevir/ritonavir plus interferon nebulization (Patient 1) GGO, ground-glass opacity. Red arrows indicate the possible lesions.

### Experienced patients treated with lopinavir/ritonavir plus interferon nebulization and switched to danoprevir/ritonavir with or without interferon nebulization

Nine treatment experienced patients were on lopinavir/ritonavir plus interferon nebulization treatment ranging from 2 to 13 days. Four Patients experienced lopinavir/ritonavir plus interferon nebulization treatment for 2 to 5 days with CT scans worsened and switched to danoprevir/ritonavir with interferon nebulization. The medical history, treatment and various examinations for one of patients, patient 6, were described in details below.

- On February 14, 2020, a man in his 40’s was admitted in to the hospital for positive RT-PCR nucleic acid test of SARS-CoV-2 in nasal swab. His family member was diagnosed with COVID-19 on February 11. Based on chest CT scans, subpleural area of the right lower lung showed GGO (Figure 3A). On admission, the patient reported a 3-day history of chest distress and fatigue. He showed no signs of fever, shortness of breath, nausea, and diarrhea. He had a history of hypertension.
- In the evening of February 14, he was given the first dose of Kaletra (lopinavir (200 mg) /ritonavir (50 mg), per tablet), 2 tablets at a time, twice per day, along with interferon nebulization.
- On February 15, the patient reported he still had chest distress. Soft thin stool was still reported and occasional cough and white sputum appeared. The patient continued lopinavir/ritonavir treatment and received rehydration supportive care.
- On February 17, 3 days after receiving lopinavir/ritonavir treatment, the lower lung showed further consolidation, along with GGO and patch-shape areas based on CT scans (Figure 3B). Therefore, in the evening of February 17, lopinavir/ritonavir treatment was stopped and the patient was switched to danoprevir boosted by ritonavir, along with interferon nebulization. On February 18, patient had a weak positive RT-PCR test for viral RNA in nasal swab.
- On February 20, 4 days after receiving danoprevir boosted by ritonavir treatment, the right lower lung showed shrunken patch-shape areas and decreased density of GGO (Figure 3C). On the same day, RT-PCR test with the nasal swab became negative and remained negative at the next day RT-PCR test.
- In the morning of February 23, the patient received the last dose of danoprevir boosted by ritonavir treatment. Since he met all four discharge standards described in Methods section, the patient was discharged from the hospital on the same day.

**Figure 3.**
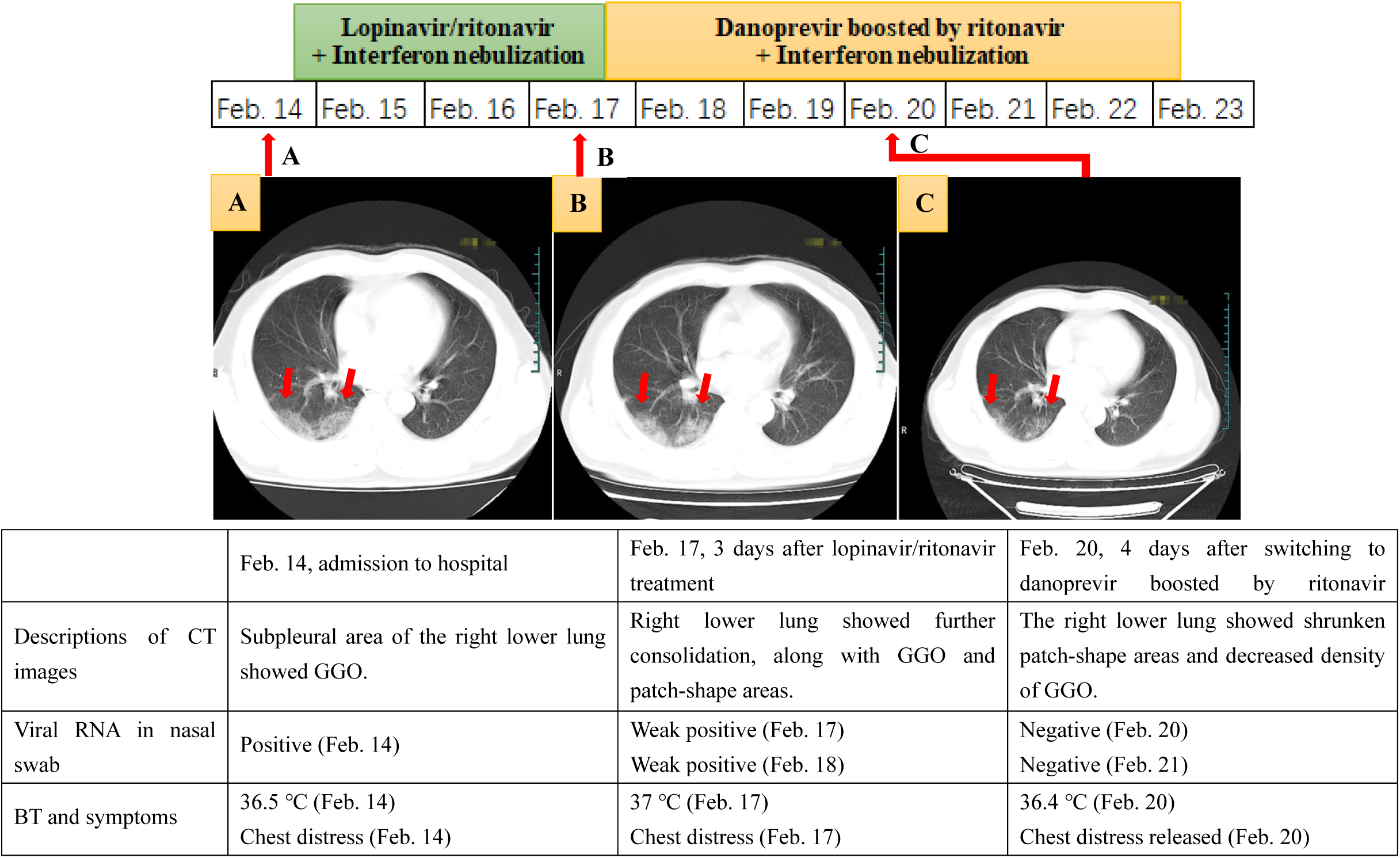
Experienced patient treated with lopinavir/ritonavir plus interferon nebulization for 3 days and switched to danoprevir/ritonavir plus interferon nebulization (Patient 6) GGO, ground-glass opacity. Red arrows indicate the possible lesions.

Five patients failed lopinavir/ritonavir plus interferon nebulization and switched to danoprevir/ritonavir without interferon nebulization. The medical history, treatment and various examinations for one of patients, patient 8, were described in details below.

- On February 9, a woman in her 40’s, who was a permanent resident in Wuhan City, was admitted to the hospital due to maximum BT 38°C and RT-PCR positive SARS-CoV-2 in nasal swab. She had occasional coughs with white sputum. The vital sign of the patient showed BT of 37.4°C and SPO2 of 99% under ambient air. The patient had half-year history of hypertension. The patient was given the first dose of lopinavir/ritonavir, 2 tablets at a time, twice per day, along with interferon nebulization in the evening of February 9.
- On February 10, based on CT scans, subpleural areas of both lower lungs and the tongue-like segment of the left lung showed GGO and patch-shape areas (Figure 4A). From February 10 to February 13, the patient showed slightly fluctuating BT (maximum 37.3°C to minimum 36.4°C), coughs with white sputum, anorexia and diarrhea with yellow thin stool. The laboratory results revealed leukopenia (3.40 ×10^9^ /L) and lymphopenia (0.7 ×10^9^ /L) (Table 3).
- On February 13, 4 days after receiving lopinavir/ritonavir treatment, GGO and patch-shape areas showed a slightly higher density (Figure 4B) compared to previous CT image on February 10.
- On February 17, 8 days after receiving lopinavir/ritonavir treatment, the patient reported improved appetite and sleeping while coughing with white sputum remained. CT scan showed that no significant changes were obat served compared to previous CT image on February 13 and the opacities at tongue-like segment increased (Figure 4C).
- On February 19, the patient received the last dose of lopinavir/ritonavir and interferon nebulization. The treatment duration of both lopinavir/ritonavir and interferon nebulization exceeded the recommendation by China National Standards [14]. As a result, both lopinavir/ritonavir and interferon nebulization were stopped.
- On February 20, she was switched to danoprevir boosted by ritonavir without interferon nebulization.
- On February 22, 3-day after receiving danoprevir/ritonavir treatment, RT-PCR test was negative in both nasal and phlegm swabs.
- On February 23, 4 days after switching to danoprevir boosted by ritonavir, the patient showed improved white blood cell count (4.31 × 10^9^ /L) (Table 3). CT scans showed GGO and patch-shape areas decreased in density compared to previous CT images on February 17, indicating absorption in both lungs (Figure 4D).
- On February 25, the patient received the last dose of danoprevir boosted by ritonavir.
- On February 26, CT scans showed the lesions in tongue-like segment of the lung were completely absorbed. Opacities in both lower lungs decreased in density compared to previous CT images (Figure 4E).
- RT-PCR tests were repeatedly negative in both nasal and phlegm swabs on February 26 and February 27.
- On February 29, the patient was discharged from the hospital since she met all four discharge standards as described in Methods section.

**Figure 4.**
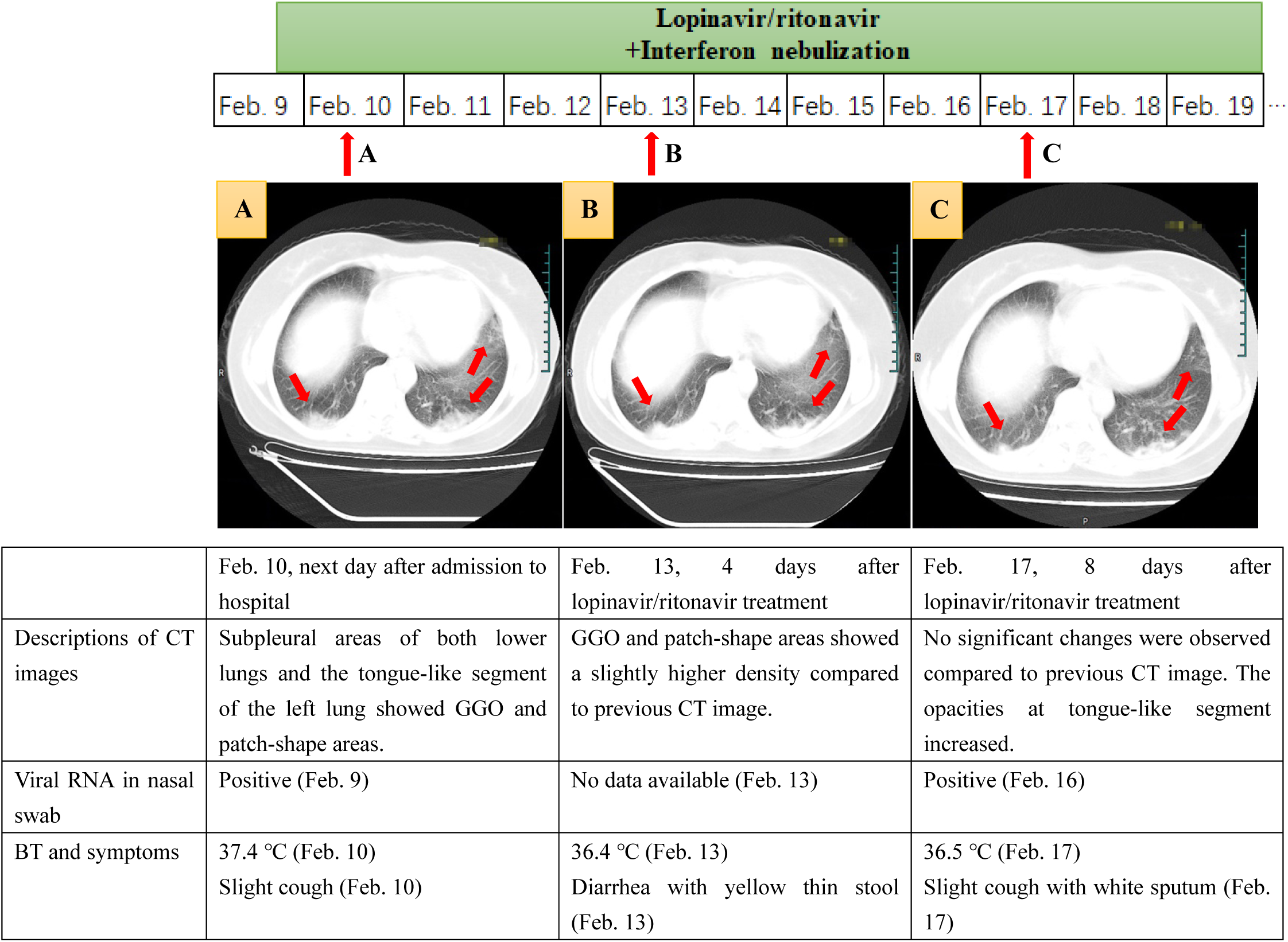
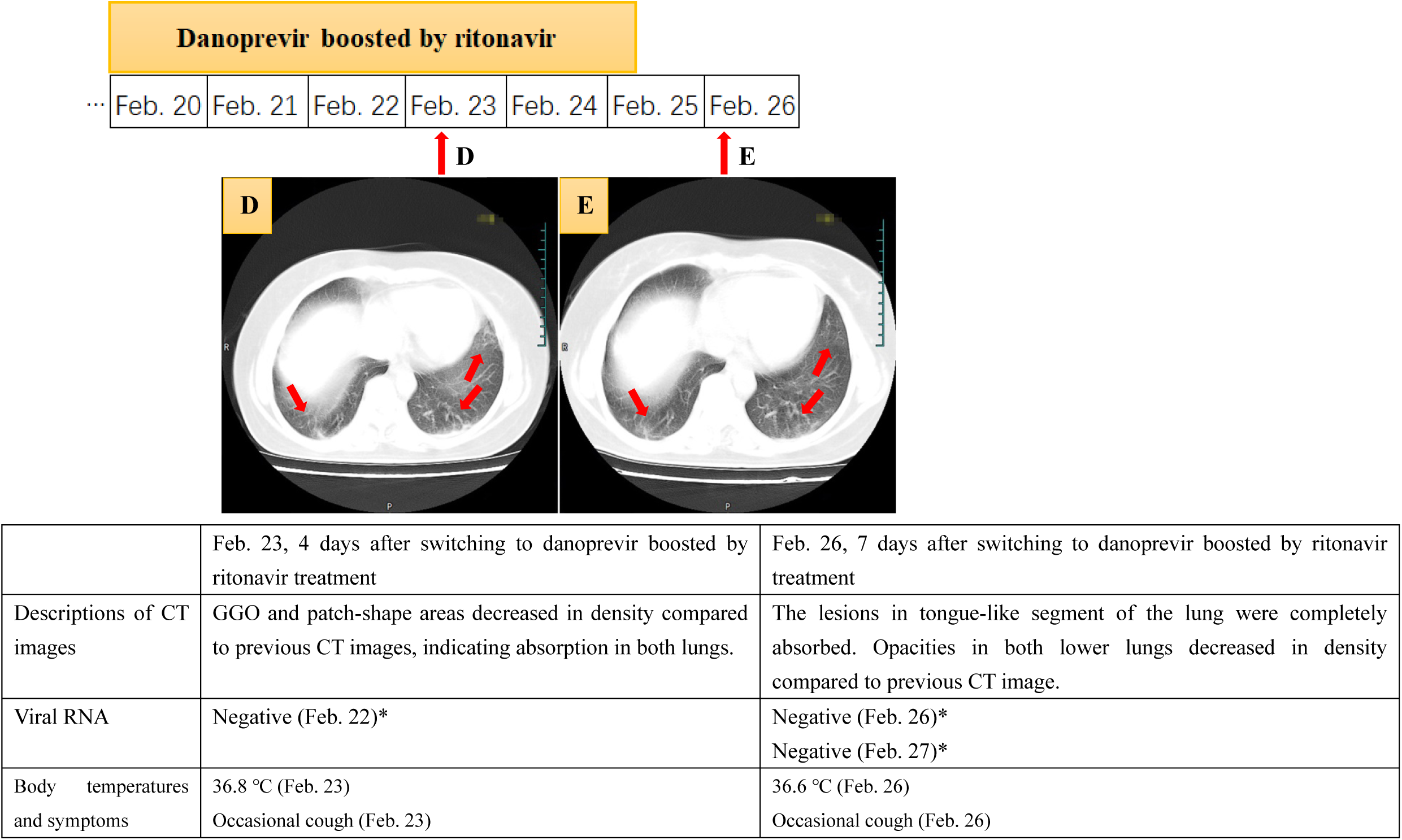
Patient failed lopinavir/ritonavir plus interferon nebulization and switched to danoprevir/ritonavir without interferon nebulization (Patient 8) GGO, ground-glass opacity. Red arrows indicate the possible lesions. * Viral RNA was undetectable in both nasal and phlegm swabs.

## Discussion

Our study for the first time demonstrated the therapeutic efficacy in moderate COVID-19 patients with the treatment of danoprevir, an HCV NS3/4A protease inhibitor marketed in China, boosted by ritonavir, a pharmacokinetic enhancer of Danoprevir and an HIV protease inhibitor. As a marketed drug, danoprevir has been demonstrated safe and well tolerated in thousands of HCV patients in China. Ritonavir is approved HIV drug and pharmacokinetic booster with more than 15-year clinical use.

Although many clinical trials involving many different drugs are ongoing in China and globally, including protease inhibitors, RNA-dependent RNA polymerase (RDRP) inhibitors, none of them has been approved for COVID-19 indication by regulatory authorities of major counties.

Around 5% of COVID-19 cases were critical with a mortality of over 50% [15], so it is essential to treat and prevent the mild or moderate cases progressing into severe or critical ones. Our report demonstrated that danoprevir boosted by ritonavir can suppress viral replication in less than one week and effectively reduce GGO and patch-shape areas. Viral nucleic acids in nasal swabs turned negative at a median of 2 days and the absorption occurred at a median of 3 days after the initiation of danoprevir/ritonavir treatment.

The mortality rate of COVID-19 patients with hypertension, cardiovascular diseases and with preexisting comorbid conditions was much higher than normal patients (6.0% to 10.5% vs 2.3%) [15]. Two patients involved in this report had a history of hypertension, but they showed a quick response with negative PCR tests in 2 days and normalized lung imaging in 3 days after the initiation of danoprevir/ritonavir treatment, respectively. This finding suggests that danoprevir/ritonavir treatment could be an effective therapy to reduce the risk of those patients.

Of 11 patients enrolled, eight patients showed significant clinical manifestations such as fever, cough and dry coughing, while three patients didn’t show significant clinical manifestations. Therefore, the most reliable indicators for treatment efficacy should be improved lung imaging via CT scans and dramatically reduced virus load through RT-PCR tests.

This study highlights the first-time successful treatment of three COVID-19 patients by HCV protease inhibitor. Nucleic acid testing results suggested 4 to 12-day treatment of danopreivir/ritonavir can effectively suppress viral replication and improve patient’s health conditions, especially for those patients with moderate COVID-19. This report also highlights the therapeutic effect of danopreivir/ritonavir treatment on both anti-viral naive and experienced patients. SARS-COV-2 is an RNA virus and thus prone to mutate under the anti-viral drug pressure. In the real world of clinic practice, patients usually experience various kinds of treatments, which could lead to drug-resistant mutations. Combination of agents that target different phases of viral life cycle should minimize the incidents of drug-resistance.

In conclusion, repurposing danoprevir, a marketed HCV protease inhibitor, represents a promising therapeutic option for COVID19 patients.

## Data Availability

The work described was original research that has not been published previously.

## Acknowledgements

We thank all the patients involved in this study; the nurses and clinical staff who are providing care for the patients; staff at the clinical laboratory of the Nineth hospital of Nanchang.

## Author contributions

HC and JW conceived and designed the study. ZZ, FG, HZ and WL contributed and performed data collection and analysis. HC, XL and JW contributed to the literature search and manuscript drafting. YC, HC and JW contributed to the planning and operation of the clinical study. All authors provided critical review of the manuscript and approved the final draft for publication.

## Abbreviations

COVID-19: coronavirus disease 2019
SARS-COV2: severe acute respiratory syndrome coronavirus-2
MERS: middle east respiratory syndrome
GGO: ground glass opacity
RT-PCR: reverse real-time PCR
HIV: human immunodeficiency virus
HCV: hepatitis C virus
3CL pro: chymotrypsin-like protease
GT: genotype
WHO: World Health Organization
SVR: sustained virologic response

